# Serum metabolite profiles are associated with the presence of advanced liver fibrosis in Chinese patients with chronic hepatitis B viral infection

**DOI:** 10.1101/2020.01.02.20016352

**Authors:** Guoxiang Xie, Xiaoning Wang, Runmin Wei, Jingye Wang, Aihua Zhao, Tianlu Chen, Yixing Wang, Hua Zhang, Zhun Xiao, Xinzhu Liu, Youping Deng, Linda Wong, Jun Panee, Cynthia Rajani, Yan Ni, Sandi Kwee, Hua Bian, Xin Gao, Ping Liu, Wei Jia

**Author notes:** Correspondence: Wei Jia, University of Hawaii Cancer Center, Honolulu, HI 96813., Ping Liu, Institute of Liver Diseases, Shuguang Hospital, Shanghai University of Traditional Chinese Medicine, 528 Zhangheng Road, Shanghai 201203, China. Authors contributed equally to this work (Guoxiang Xie, Xiaoning Wang, Runmin Wei, Jingye Wang). **Author contributions:** W.J. was principal investigator of this study and designed the study. P.L. organized the patient recruitment and provided biospecimens for this study. G.X.X. drafted the manuscript. W.J., G.X.X., X.N.W., R.M.W., J.Y.W., A.H.Z., L.W., J.P., C.R., S.K. H.B., and X.G. critically revised the manuscript. R.M.W., J.Y.W., G.X.X., X.N.W., A.H.Z., and Y.N. performed data analysis. X.N.W., Y.X.W., H.Z., Z.X., and X.Z.L. were responsible for clinical data collection. G.X.X., X.N.W., and A.H.Z. performed the metabolomics analysis.

## Abstract

**Background & Aims:** Accurate and noninvasive diagnosis and staging of liver fibrosis is essential for effective clinical management of chronic liver disease (CLD). We aimed to identify serum metabolite markers that reliably predict the stage of fibrosis in CLD patients.

**Methods:** We quantitatively profiled serum metabolites of participants in 2 independent cohorts. Based on the metabolomics data from Cohort 1 (504 HBV associated liver fibrosis patients and 502 normal controls, NC), we selected a panel of 4 predictive metabolite markers. Consequently, we constructed 3 machine learning models with the 4 metabolite markers using random forest (RF), to differentiate CLD patients from normal controls (NC), to differentiate cirrhosis patients from fibrosis patients, and to differentiate advanced fibrosis from early fibrosis, respectively.

**Results:** The panel of 4 metabolite markers consisted of taurocholate, tyrosine, valine, and linoelaidic acid. The RF models of the metabolite panel demonstrated the strongest stratification ability in Cohort 1 to diagnose CLD patients from NC (area under the receiver operating characteristic curve (AUROC) = 0.997 and the precision-recall curve (AUPR) = 0.994), to differentiate fibrosis from cirrhosis (0.941, 0.870), and to stage liver fibrosis (0.918, 0.892). The diagnostic accuracy of the models was further validated in an independent Cohort 2 consisting of 300 CLD patients with chronic HBV infection and 90 NC. The AUCs of the models were consistently higher than APRI, FIB-4 and AST/ALT ratio, with both greater sensitivity and specificity.

**Conclusion:** Our study showed that this 4-metabolite panel has potential usefulness in clinical assessments of CLD progression.

## INTRODUCTION

Liver fibrosis is a wound-healing response to damage caused by chronic liver disease (CLD) [1]. Liver fibrosis can progress to cirrhosis over years or decades [2], and results in liver function decline and increased risk of hepatocellular carcinoma (HCC). Liver biopsy has been the gold standard for evaluating the presence and degree of liver fibrosis, but its clinical application is limited by inherent limitations such as invasiveness, sampling errors, and intra- and inter-observer variability [3]. Recent studies indicated that liver fibrosis could be reversed [1], creating the need for less invasive clinical tools to monitor and assess the responses of CLD patients to treatments. A number of scoring systems, such as the FibroTest,[4] the aspartate transaminase/alanine transaminase (AST/ALT) ratio [5], the AST/Platelet Ratio Index (APRI) [6], FIB-4 (patient age, AST, ALT, and platelet) [7], *Wisteria floribunda* agglutinin-positive Mac-2 binding protein (WFA^+^-M2BP) [8] and machine learning-based clinical predictive models [9] have recently been used to stage CLD and predict the development of liver fibrosis and cirrhosis. Imaging techniques, such as computed tomography, magnetic resonance imaging [10], and two recently approved ultrasound-based systems, shear wave elastography and transient elastography (FibroScan) [11], have also been used clinically to assess the degree of liver fibrosis. However, these imaging modalities have limited accuracy in some patients, such as those with ascites, elevated central venous pressure, and obesity [12].

Developing noninvasive, accurate, and reliable markers to assess the severity and progression of liver fibrosis in CLD patients has become increasingly important for treatment decisions, for continuous monitoring of patients who have mild liver disease and are not under treatment [13], and for risk stratification and longitudinal followup in clinical trials.

Alterations of bile acids (BAs) [13-19], free fatty acids (FFAs) [20], and amino acids (AAs) [21, 22] are closely associated with CLD regardless of etiology. However, the relationship between serum AAs, BAs, and FFAs and the stages of liver fibrosis have not been thoroughly investigated. The aim of this study was to identify serum metabolite markers that reliably predict the stage of fibrosis in CLD patients with chronic hepatitis B virus (HBV) infection, a leading cause of CLD worldwide. We used a targeted metabolomics approach to quantify serum BAs, AAs, and FFAs in 1,006 participants in Cohort 1 (504 biopsy-proven fibrosis and cirrhosis CLD patients with chronic HBV infection and 502 normal controls, NC), and selected four predictive metabolite markers to construct three machine learning models using random forest (RF). Model 1 diagnosed CLD patients from NC, Model 2 differentiated cirrhosis patients from fibrosis patients, and Model 3 differentiated advanced fibrosis and early fibrosis patients. The diagnostic accuracy of the three models was further validated in an independent cohort consisting of 300 HBV-CLD patients and 90 NC.

## MATERIALS AND METHODS

### Study design and participants

Two data sets were enrolled in this study. Cohort 1 was recruited between April 2013 and June 2015 at Shuguang Hospital Affiliated to Shanghai University of Traditional Chinese Medicine, consisted of 1,006 participants, including 504 CLD patients with chronic HBV infection and 502 NC as our training cohort to identify serum metabolite markers and establish predictive models (Table 1). More detailed inclusion and exclusion criteria can be found in the Supporting Information.

**Table 1:**
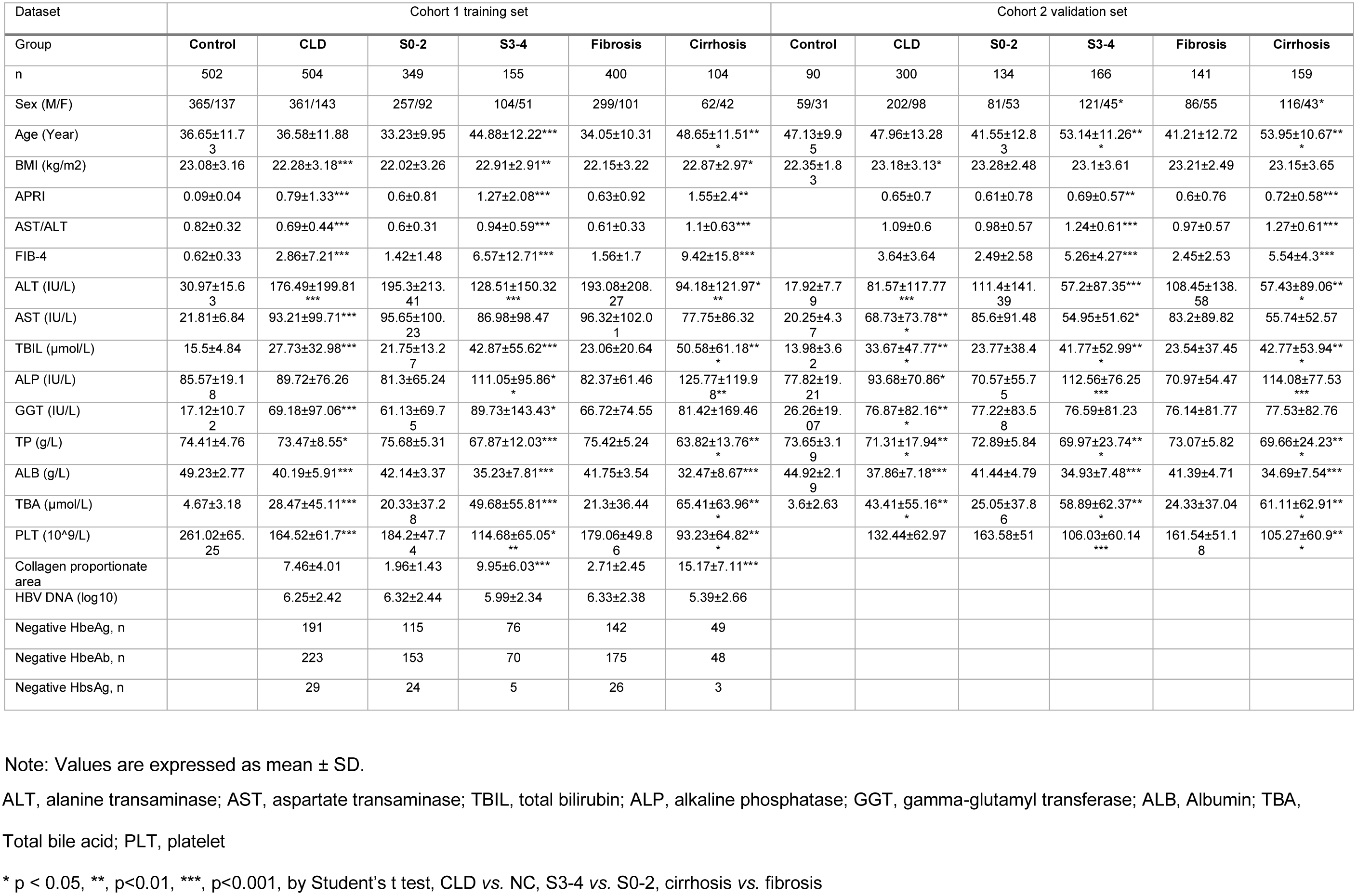
Demographic and clinical data of patients with CLD and NC in Cohort 1 (training set) and Cohorts 2 (validation set)

Cohort 2, recruited between December 2016 and December 2017 at Xiamen Hospital of Traditional Chinese Medicine, consisted of 300 CLD patients with chronic HBV infection and 90 NC. Data obtained from Cohort 2 were used as a validation set to further verify the performance of the models established from the Cohort 1. Detailed information about this cohort can be found in the Supporting Information. Sample size was not determined by statistical methods and was comparable to other studies in the field [4-8, 21, 22].

In this study, the diagnosis and the sample collection were performed using exactly the same protocols to avoid “external” influences. The samples were provided to lab staffs blind samples with respect to patient identity and other clinical information.

The study was approved by the institutional review board of each hospital. All participants provided written informed consent.

#### Liver biopsy

Detailed information is provided in the Supporting Materials and Methods.

#### Histological assessment of liver injury

Detailed information is provided in the Supporting Materials and Methods and Fig. S7.

#### Collagen proportionate area using digital image analysis (DIA)

Detailed information is provided in the Supporting Materials and Methods and Fig. S8.

#### Serum sample collection, blood clinical marker measurement, and metabolomics analysis

The procedure and analysis was performed as described in the Supporting Materials and Methods.

### Classification Performance Evaluation

ROC curve is a plot of the true positive rate (sensitivity/recall) against the false positive rate (1-specificity) at different cutoffs of a binary classifier. AUROC measures the area under the ROC curves and a higher value of AUROC suggests better classification performances while an AUROC of 0.5 represents the random guess. The PR curve demonstrates the relationship between positive predictive values (precision) and true positive rate (sensitivity/recall), and a higher value of AUPR indicates better diagnostic capacity of the model. PR curves are usually preferable for evaluating unbalanced data compared to ROC curves. NRI and IDI were also used for the evaluation of prediction improvement. We compared RF models to existing clinical indices by splitting the continuous risk scores into ten equal risk intervals (default). We used the R software version 3.2.3 for data analysis and the “PRROC” R package for binary ROC and PR curves[23], the “pROC” package for calculating the specificities and sensitivities of classifiers[24], and the “PredictABEL” package for NRI and IDI calculation[25].

### Feature Selection and Method Comparison

Quantitative variables were expressed as mean ± SD for clinical parameters and median (25% quantile, 75% quantile) of log10 transformed concentration for metabolites. Categorical variables were expressed as percentages. The univariate analysis (Wilcoxon rank-sum test) was carried out to identify the variables that were significantly different between CLD patients and NC, between fibrosis and cirrhosis (S0-3 vs. S4), and among CLD patients at different fibrotic stages (early stage fibrosis (S0-2) vs. advanced stage fibrosis (S3-4)).

For differential metabolites with p < 0.001 across all univariate analyses were used in two machine learning methods, LASSO[26] and RF[27] to further select markers for the three classifications listed above. Data were log and z-score transformed before being fed into LASSO to ensure that the coefficients were comparable with each other. The regularization parameter lambda of LASSO was determined using 10-fold CV. The RF model used 500 decision trees. We ranked the metabolites according to their LASSO non-zero coefficients and RF mean decrease of accuracy, and kept the intersection of top 5 LASSO and RF metabolites in the three classifications. Considering the overlaps of the second and the third classification tasks, we further selected the intersecting variables of these two situations and then, merged with variables selected from the first situation to construct our final metabolite markers (Fig. 2).

**Figure 2.**
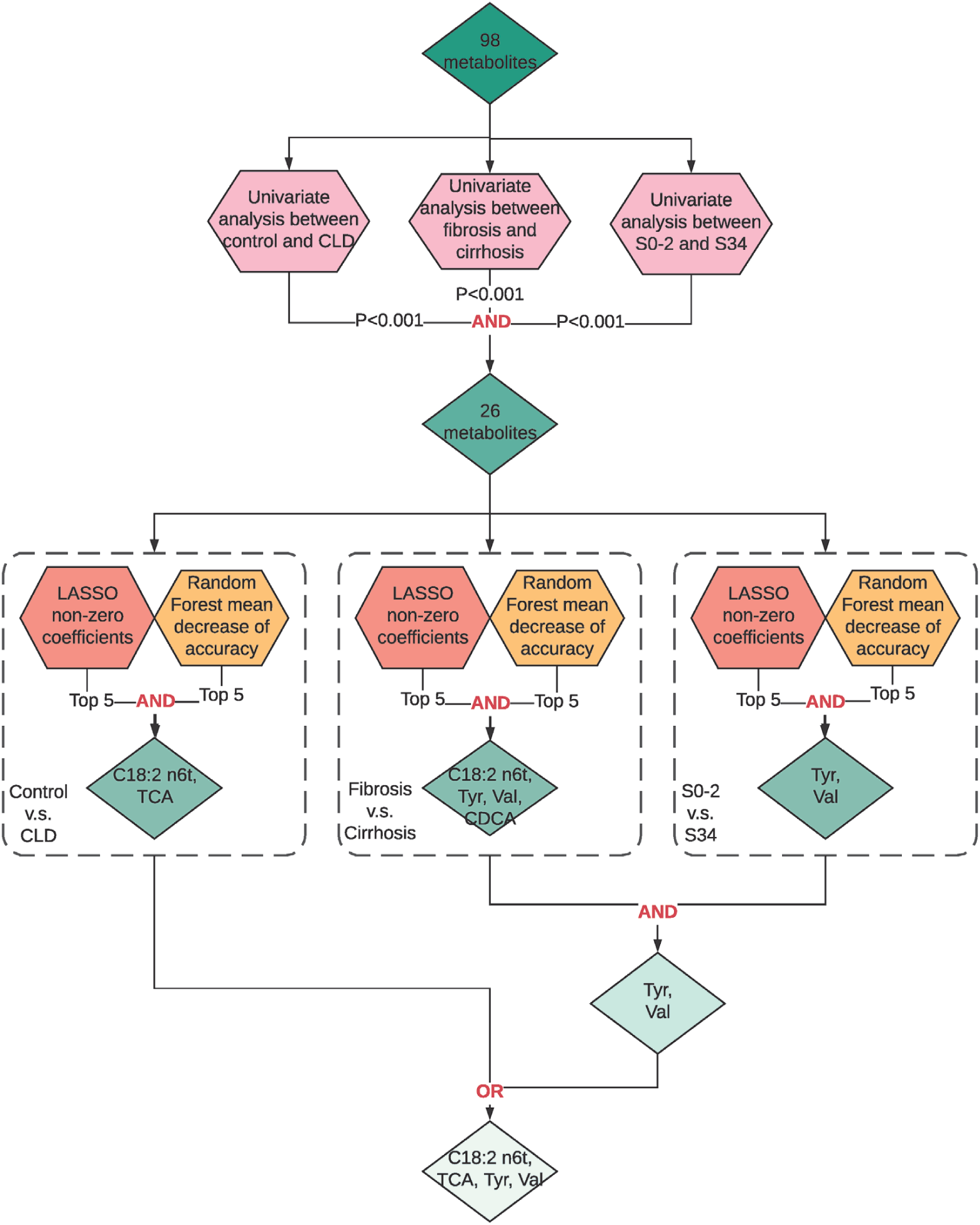
Workflow chart of feature selection. For a total of 98 metabolites (including AAs, BAs and FFAs), univariate analyses (Wilcoxon rank-sum test) were employed for three clinical aims (aim-1: CLD vs. NC, aim-2: Cirrhosis vs. Fibrosis, aim-3: Early fibrosis vs. Advanced fibrosis). 26 metabolites with P < 0.001 in all three clinical aims were selected and fed into LASSO and Random Forest algorithms for three aims. The overlap of top-5 LASSO non-zero coefficients and top-5 important variables from Random Forest (by mean decrease of accuracy) were selected. For aim-2 and aim-3, we selected the overlapped variables and combined with variables selected from aim-1 to yield the final panel four metabolites. Note: “OR” means the union of two sets, “AND” means the intersection of two or more sets.

To identify an appropriate classification method, we introduced two linear models, i.e., logistic regression (LR), linear discriminant analysis (LDA) and one decision tree-based ensemble model, i.e., RF, for the classifier construction for the markers we selected. For RF, we used 500 decision trees and two candidate variables at each split. For LDA, the tolerance parameter was set to 1.0E-4 (default). We applied 10-fold CV on the training set (Cohort 1) to compare the classification performances of these four models and three established fibrosis markers, i.e., APRI, AST/ALT ratio, and FIB-4. AUROC and AUPR were recorded at each internal validation set in CV. We used R packages “randomForest”, “glmnet” and “MASS” for RF, LASSO and LDA constructions respectively [28, 29].

### Predictive Model Construction and Validation

We trained the final RF models for different classification objectives using the training data (Cohort 1), with Model 1 differentiating CLD and NC, Model 2 differentiating fibrosis and cirrhosis, and Model 3 differentiating early and advanced stages of liver fibrosis. A total of 500 decision trees were included in a single RF model with two variables randomly sampled as candidates at each split. We re-balanced the sample size for different groups at each bootstrap resampling step for Models 2 and 3 considering the unbalanced samples.[30]

In RF, each decision tree was fitted on the bootstrap samples and tested on the untouched OOB-samples. Thus, the OOB-predictions provided unbiased estimates of how the RF model performed on the training data and were used for the evaluation on Cohort 1. We further validated our mark panel-based RF models in the independent validation data sets from Cohort 2, and compared results with the established fibrosis markers, AST/ALT ratio, APRI and FIB-4. ROC and PR curves were drawn and AUROC and AUPR values, respectively, were calculated to evaluate their diagnostic performances. Optimal cut-offs were selected to maximize the sum of sensitivity and specificity for the RF model. For APRI, FIB-4 and AST/ALT, predefined cut-offs were used (1.0 and 2.0 for APRI to distinguish fibrosis and cirrhosis,[6] 1.45 and 3.25 for FIB-4 to distinguish S0-2 and S3-4,[7] and 0.8 and 1.0 for AST/ALT to distinguish S0-2 and S3-4[5, 31]). Bootstrap resampling (1,000 times) was conducted to calculate 95% confidence intervals (CIs) of AUCs for all binary classifiers. A comparison of the AUROC of our biomarker panel vs. FIB-4, AST/ALT, or APRI was performed using DeLong’s test. The significance level was adjusted for multiple testing according to the Benjamin and Hochberg procedure.[32] Log and z-score transformed data were also used for constructing heatmaps. The R packages “ggplot2” and “cowplot” were used for data visualization and multiple plots arrangement.

We further derived an RF risk score for each participant based on the marker panel and logit function of the predicted probability (Prob.) of the RF model for corresponding classification objective:

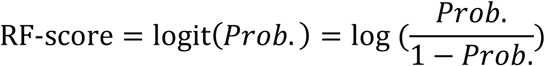

F1 scores were then calculated at the predefined cutoffs using following formula:

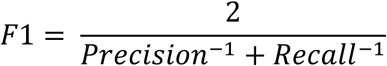

To determine whether the RF score could independently predict the fibrosis staging in the presence of other potential confounding factors, we applied logistic regression on the RF-score to differentiate cirrhosis from fibrosis as well as discriminate early and advance fibrosis while adjusting for HBV DNA levels, the degree of necro-inflammation, HBeAb status, HBeAg status, liver function tests (i.e., PT, ALB, DBIL, IBIL), platelets, BMI, and medication (entecavir) use.

### Multi-group Classification of S0-2 vs. S3 vs. S4

We built a new RF model based on our metabolite marker panel and applied multinomial regressions to APRI, AST/ALT, and FIB-4 separately to differentiate S0-2 vs. S3 vs. S4 in Cohort 1. Then, we compared and validated these multi-group classifiers on both Cohort 1 and Cohort 2 data sets using micro-average ROC and PR curves. Micro-average ROC and PR curves were calculated by stacking binary classification results from each group together to generate a concatenated binary classification result.[33] We then calculated AUROC and AUPR with 95% CIs using 100 times bootstrap resampling. We used the “multiROC” R package for calculating the micro-average AUROC and AUPR as well as for plotting[34].

### Code availability

R, GraphPad, and in-house metabolomic processing pipeline were used for the data analyses presented in this study. We used GraphPad Prism 7, R software version 3.2.3 with packages including PRROC, pROC, PredictABEL, randomForest, glmnet, MASS, ggplot2, cowplot and multiROC. The metabolomics data were processed and analyzed by TargetLynx (Waters). The computer code used to generate the results reported in this study is available from the authors upon request.

### Data availability statement

All the data supporting the findings of this study are available within the article and its Supplementary Information files or from the corresponding author upon reasonable request.

## RESULTS

### Characteristics of the participants

Two independent cohorts were studied (Fig. 1). CLD groups were staged and assigned according to the results of their liver biopsy. Cohort 1 consisted of 1,006 participants (502 NC and 504 biopsy-proven HBV-CLD patients (400 with liver fibrosis (S0-3) and 104 with cirrhosis (S4); or 349 with early stage fibrosis (S0-2) and 155 with advanced stage fibrosis (S3-4)). Cohort 2 consisted of 390 participants (90 NC and 300 biopsy-proven CLD patients comprising 141 with fibrosis and 159 with cirrhosis, or 134 with early stage fibrosis (S0-2) and 166 with advanced stage fibrosis (S3-4). Models established from Cohort 1 were validated in Cohort 2 (Models 1, 2 and 3). The cohort stratification and major demographic and clinical characteristics are shown in Table 1. More detailed clinical data are provided in Table S1.

**Figure 1.**
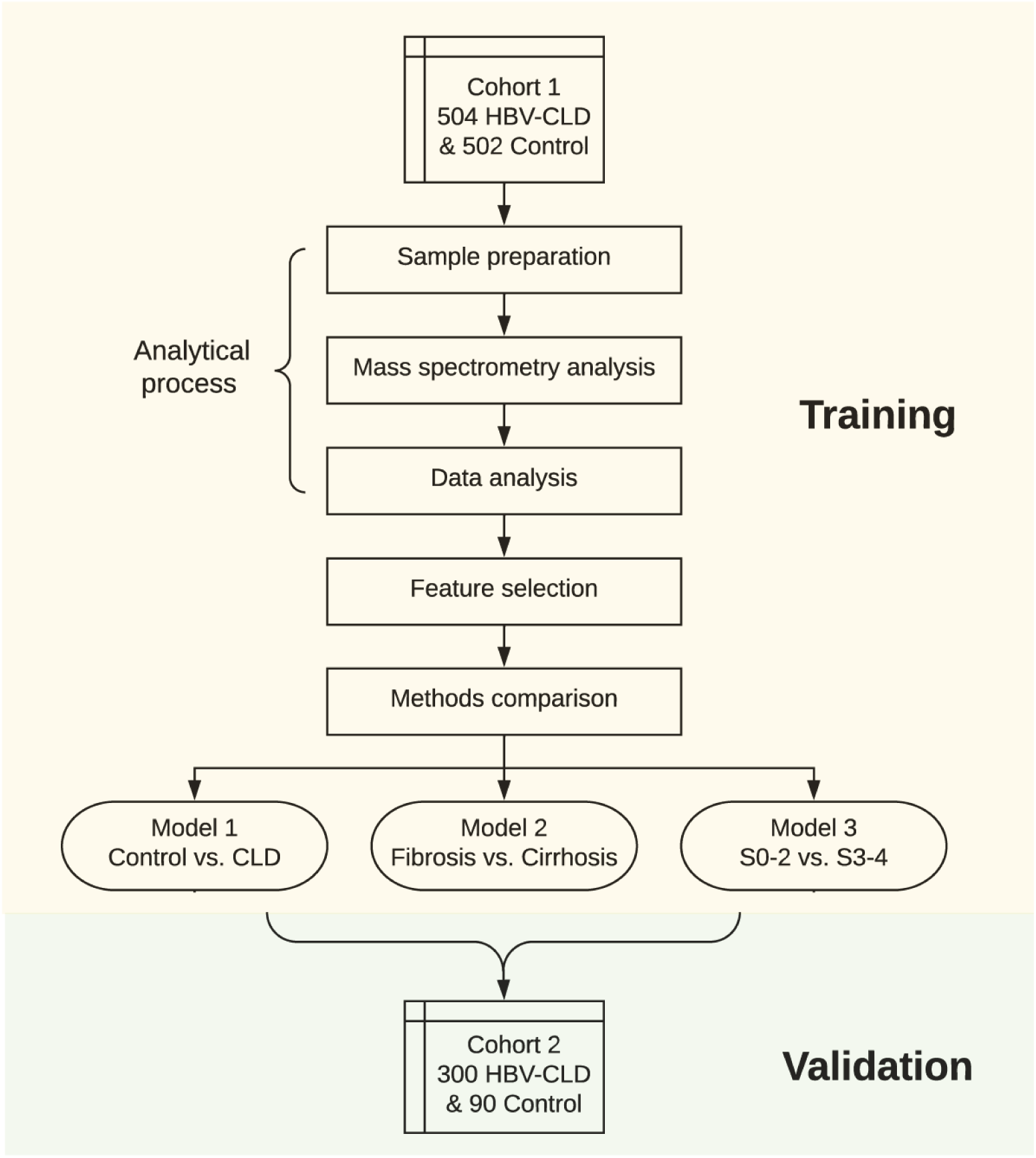
Study design. Serum metabolites were quantified in Cohort 1 (504 biopsy-proven HBV-CLD patients and 502 NC) and were used to identify candidate markers. After data analysis and feature selection, four metabolites were selected to compose our marker panel. Different machine models and clinical indices were compared using 10-fold cross validation. Three RF models were constructed to diagnose CLD from NC (Model 1), differentiate fibrosis vs. cirrhosis (Model 2) and grade early fibrosis vs. advanced fibrosis (Model 3) in Cohort 1. These three were further validated in the independent HBV Cohort 2.

### Quantification of metabolites in serum

Using targeted metabolomic protocols establlished in our lab,[35-37] we quantified the concentrations of 98 metabolites, including 24 BAs, 42 FFAs and 32 AAs, in the sera of all participants (Table S2). These metabolites were used for the subsequent metabolite marker selection.

### Serum metabolite marker selection

From the 98 serum metabolites, we identified 26 differential metabolites in three classification situations (i.e., to diagnose CLD patients from NC, to differentiate fibrosis from cirrhosis, and to differentiate advanced fibrosis from early fibrosis) using univariate analysis (Wilcoxon rank sum test, P<0.001). The 26 statistically significant metabolites were then entered into least absolute shrinkage and selection operator (LASSO)[26] and random forest (RF)[27]. According to the rank of LASSO non-zero coefficients and RF mean decrease of accuracy, four metabolite markers were selected, which included one FFA, linoelaidic acid (C18:2 n6t), one BA, taurocholate (TCA), and two AAs, tyrosine (Tyr) and valine (Val) (Fig. 2). The principal component analysis (PCA) of these four metabolite markers also showed a clear separation between CLD patients and NC (Fig. S1). We also derived one ratio, the Tyr/Val ratio to further improve the classification performances while also including one extra accessible risk factor, age, to our panel for the differentiation of fibrosis and cirrhosis and the staging of fibrosis. Correlations of the four metabolites with fibrosis stage, necro-inflammation, CPA, AST, ALT, AST/ALT ratio, PLT, FIB-4, and APRI were assessed using Spearman correlation analysis (Fig. S2). The four metabolite markers (including the Tyr/Val ratio) all significantly correlated with fibrosis stage (ρ=0.38 for TCA, ρ=0.50 for Tyr, ρ=0.53 for Tyr/Val ratio and ρ=0.23 for C18:2 n6t) using Spearman’s correlation analysis. In addition, we found our metabolite markers showed stronger associations with the fibrosis stage than the previously used clinical indices.

To determine an appropriate classification model, we applied 10-fold cross-validation (CV) on Cohort 1 to compare the classification performances of RF models and two linear models (i.e., logistic regression (LR), linear discriminant analysis (LDA)) as well as the clinical indices, APRI, AST/ALT ratio, and FIB-4. The CV-area under the receiver operating characteristic curve (CV-AUROC) and area under the precision-recall curve (CV-AUPR) were employed as the evaluation metrics. We found that, to differentiate CLD from control, APRI, LR, LDA and RF had the highest AUROCs and AUPR, while RF demonstrated the most robust classification performance (Fig. S3a). For the differentiation of fibrosis and cirrhosis and S0-2 vs. S3-4, RF outperformed other methods with the highest CV-AUROC and CV-AUPR overall (Figs. S3b, c). PCA scores plot showed linearly separable discrimination between the most CLD and control subjects (Fig. S1), thus linear models could achieve good classification performances. However, for a situation where there is more extensive overlapping of groups (Fig. S4), the decision tree-based ensemble learning algorithm, RF, achieved improved classification performances compared to other methods (Fig. S3).

### Model 1: Differentiating CLD patients from NC

The concentration of linoelaidic acid (C18:2 n6t) was significantly higher in the control group than in the CLD group, conversely the levels of TCA, and Tyr, and Tyr/Val ratio were higher in the CLD group than in the control group (Figs. 3a, b).

**Figure 3.**
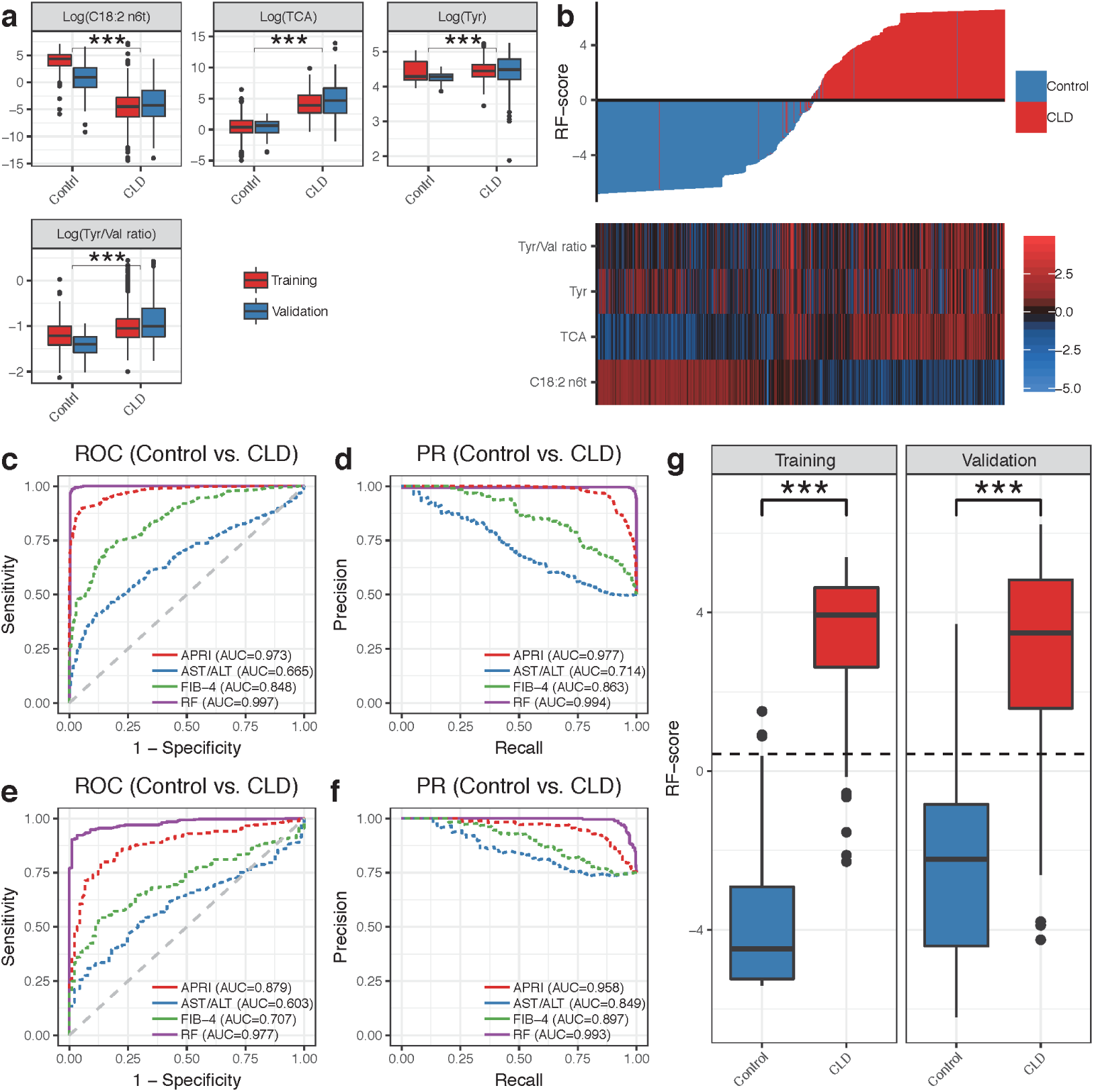
Metabolite marker panel and Model 1 for CLD with chronic HBV infection diagnosis. (a) Comparison of the four markers between CLD patients and NC in Cohorts 1 and 2. (b) Waterfall plot of RF-score and corresponding heatmap for the four markers in all data sets. (c) ROC curves of Model 1 (RF model constructed with four markers), APRI, AST/ALT and FIB-4 in Cohort 1. (d) PR curves of Model 1, APRI, AST/ALT and FIB-4 in Cohort 1. (e) ROC curves of Model 1, APRI, AST/ALT and FIB-4 in Cohort 2 validation set. (f) PR curves of Model 1, APRI, AST/ALT and FIB-4 in Cohort 2 validation set. (g) The diagnosis RF-score in NC and CLD patients in training and validation sets. *** p < 0.001, Wilcoxon rank sum test. The optimal cut-off value of the RF-score was 0.434.

Model 1 was constructed using an RF model that utilized these four metabolite markers, to differentiate CLD patients from NC in Cohort 1. Out-of-bag (OOB) estimates were employed for the RF model evaluations. Model 1 showed an AUROC of 0.997 (0.993-1.000) and AUPR of 0.994 (0.998-1.000) (Figs. 3c, d) which was significantly higher than the APRI (AUROC=0.973, p<0.001), FIB-4 (AUROC=0.848, p<0.001) and AST/ALT ratio (AUROC=0.665, p<0.001) (Table 2). An example decision tree from the RF model is shown in Fig. S5a, where we observed that the lower concentration of C18:2 n6t and the higher concentration of TCA would lead to higher risk of CLD.

**Table 2:** Results for measurement of the metabolite marker panel, APRI, FIB-4, and AST/ALT ratio in the prediction of liver fibrosis

|  | Cohort 1 training set |  |  | Cohort 2 validation set |  |  |
| --- | --- | --- | --- | --- | --- | --- |
|  | CLD vs controls | Fibrosis vs Cirrhosis | S0-2 vs S34 | CLD vs controls | Fibrosis vs Cirrhosis | S0-2 vs S34 |
| **Metabolite marker panel** |  |  |  |  |  |  |
| AUROC (95% CI)<sup>#</sup> | 0.997 (0.993-1) | 0.941 (0.914-0.964) | 0.918 (0.889-0.946) | 0.977 (0.963-0.988) | 0.844 (0.797-0.884) | 0.807 (0.756-0.852) |
| AUPR (95% CI) | 0.994 (0.986-1) | 0.87 (0.824-0.913) | 0.892 (0.854-0.925) | 0.993 (0.989-0.997) | 0.827 (0.761-0.884) | 0.817 (0.764-0.866) |
| Cutoff value (Sensitivity (%)/Specificity (%)/F1(%))<sup>†</sup> | 0.434<br>(98.4/99/98.7) | 0.01 (87/90.4/78.4) | -0.115<br>(86.7/90.5/84.6) | 0.434<br>(92.2/94.4/95.2) | 0.01<br>(71.8/81.6/73.3) | -0.115 (72.9/76.1/71.8) |
| **FIB-4** |  |  |  |  |  |  |
| AUROC (95% CI) | 0.848 (0.823-0.87) | 0.869 (0.829-0.906) | 0.802 (0.762-0.844) | 0.707 (0.652-0.762) | 0.758 (0.692-0.815) | 0.739 (0.68-0.798) |
| AUPR (95% CI) | 0.863 (0.84-0.883) | 0.725 (0.657-0.79) | 0.707 (0.651-0.761) | 0.897 (0.873-0.918) | 0.726 (0.657-0.794) | 0.726 (0.66-0.795) |
| Cutoff value-1 (Sensitivity (%)/Specificity (%)/F1(%))<sup>*</sup> |  |  | 1.45 (68/73.6/62) |  |  | 1.45 (81.5/42.5/68.1) |
| Cutoff value-2 (Sensitivity (%)/Specificity (%)/F1(%))<sup>*</sup> |  |  | 3.25 (44.2/93.4/56.3) |  |  | 3.25 (57/81.3/65.5) |
| **APRI** |  |  |  |  |  |  |
| AUROC (95% CI) | 0.973 (0.965-0.981) | 0.698 (0.644-0.752) | 0.647 (0.595-0.698) | 0.879 (0.841-0.915) | 0.608 (0.534-0.671) | 0.595 (0.529-0.669) |
| AUPR (95% CI) | 0.977 (0.969-0.983) | 0.416 (0.345-0.497) | 0.492 (0.434-0.554) | 0.958 (0.942-0.972) | 0.53 (0.474-0.605) | 0.542 (0.488-0.614) |
| Cutoff value-1 (Sensitivity (%)/Specificity (%)/F1(%))<sup>**</sup> |  | 1 (33.9/86.2/37.1) |  |  | 1 (22.7/84.4/32.4) |  |
| Cutoff value-2 (Sensitivity (%)/Specificity (%)/F1(%))<sup>**</sup> |  | 2 (18.3/94.6/26.6) |  |  | 2 (3.9/94.3/8.5) |  |
| **AST/ALT** |  |  |  |  |  |  |
| AUROC (95% CI) | 0.665 (0.631-0.697) | 0.815 (0.766-0.862) | 0.714 (0.668-0.759) | 0.603 (0.54-0.657) | 0.684 (0.619-0.75) | 0.667 (0.597-0.728) |
| AUPR (95% CI) | 0.714 (0.685-0.747) | 0.579 (0.496-0.674) | 0.582 (0.516-0.654) | 0.849 (0.819-0.875) | 0.641 (0.573-0.721) | 0.648 (0.583-0.727) |
| Cutoff value-1 (Sensitivity (%)/Specificity (%)/F1(%))<sup>***</sup> |  |  | 0.8 (48.1/84.8/54.2) |  |  | 0.8 (78.5/42.5/66.7) |
| Cutoff value-2 (Sensitivity (%)/Specificity (%)/F1(%))<sup>***</sup> |  |  | 1 (33.1/92.8/45.1) |  |  | 1 (65.9/59/63.8) |
| **Comparison of AUROC** |  |  |  |  |  |  |
| Metabolite marker panel versus FIB-4 **** | p<0.001 | p<0.001 | p<0.001 | p<0.001 | p<0.001 | 0.01 |
| Metabolite marker panel versus APRI **** | p<0.001 | p<0.001 | p<0.001 | p<0.001 | p<0.001 | p<0.001 |
| Metabolite marker panel versus AST/ALT **** | p<0.001 | p<0.001 | p<0.001 | p<0.001 | p<0.001 | p<0.001 |

Based on the OOB predicted probabilities, we calculated a diagnostic RF-score for Model 1 using the logit function. The waterfall plot showed a clear ascending trend of RF-scores from NC (lower RF-scores) to CLD patients (higher RF scores) along with the differentiation trend shown in the heatmap of the four markers (Fig. 3b). We observed significant differences in the RF-score between both groups in Cohort 1 (p<0.001, Fig. 3g), yielding a sensitivity of 98.4% and specificity of 99% for CLD patients in the training set at a cutoff value of 0.434 (Table 2). The sensitivity and specificity of our RF model were superior to those of AST/ALT ratio, APRI, and FIB-4 for differentiating CLD patients from NC using the optimal cutoffs generated in Cohort 1 using the Youden index (Table S4).

### Model 2: Differentiating cirrhosis from fibrosis among CLD patients

The discriminant prediction model was constructed using an RF model employing the four metabolite markers along with age to differentiate CLD patients with cirrhosis from those without cirrhosis in Cohort 1. This model demonstrated an AUROC of 0.941 (0.914-0.964) and AUPR of 0.87 (0.824-0.913) (Figs. 4a, b) based on OOB predictions. These results were better than those of the APRI (AUROC=0.698, p<0.001), AST/ALT (AUROC=0.815, p<0.001) and FIB-4 (AUROC=0.869, p<0.001) (Table 2). We showed an example decision tree for Model 2 in Fig. S5b, and we found that the higher the Tyr/Val ratio, Tyr and C18:2 n6t, the higher the risk of CLD with cirrhosis.

**Figure 4.**
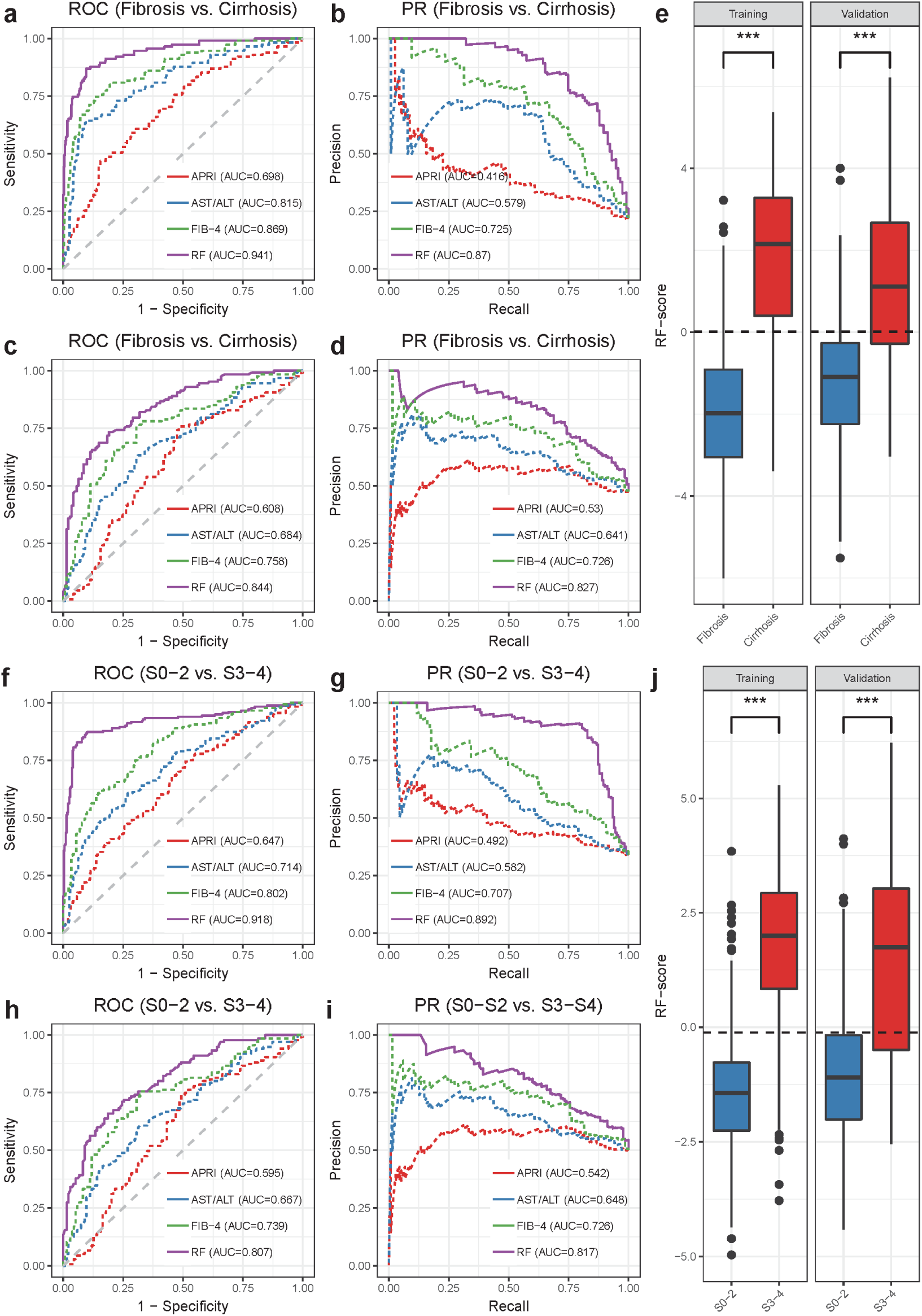
Model 2 for differentiating fibrosis vs. cirrhosis and Model 3 for differentiating early fibrosis vs. advanced fibrosis in CLD patients with chronic HBV infection. (a) ROC curves of Model 2 (RF model constructed with four metabolite markers and age), APRI, AST/ALT, and FIB-4 in Cohort 1. (b) PR curves of Model 2, APRI, AST/ALT, and FIB-4 in Cohort 1. (c) ROC curves for Model 2, APRI, AST/ALT, and FIB-4 for the Cohort 2 validation set. (d) PR curves for Model 2, APRI, AST/ALT, and FIB-4 for the Cohort 2 validation set. (e) The RF-score in CLD patients with fibrosis and cirrhosis in the HBV training, validation sets. The optimal cut-off value of the RF-score was 0.01. (f) ROC curves of Model 3 (RF model constructed with four metabolite markers and age), APRI, AST/ALT, and FIB-4 in Cohort 1. (b) PR curves of Model 3, APRI, AST/ALT, and FIB-4 in Cohort 1. (c) ROC curves for Model 3, APRI, AST/ALT, and FIB-4 for the Cohort 2 validation set. (d) PR curves for Model 3, APRI, AST/ALT, and FIB-4 for the Cohort 2 validation set. (e) The RF-score in CLD patients with S0-2 and S3-4 in the HBV training, validation sets. The optimal cut-off value of the RF-score was −0.115. *** p < 0.001, Wilcoxon rank sum test.

The Model 2 RF-score differentiated CLD patients with cirrhosis from fibrosis in Cohort 1 (p<0.001) (Fig. 4e). The constructed model yielded a sensitivity of 87.0% and specificity of 90.4% in the Cohort 1 data set at a cutoff value of 0.01 (Table 2). The RF-scores remained significant with a coefficient of 0.755 (p<0.001) after adjusting for HBV DNA levels, degree of necro-inflammation, HBeAb status, HBeAg status, body mass index (BMI), platelets (PLT), liver function tests (i.e., prothrombin time (PT), albumin (ALB), direct bilirubin (DBIL), indirect bilirubin (IBIL)), and medication (Entecavir) (Table S3). The accuracy of our RF model was superior to those of AST/ALT ratio, APRI, and FIB-4 (Table 2 and Table S4).

### Model 3: Differentiating advanced fibrosis from early fibrosis among CLD patients

In this study, fibrosis stages 0-2 were defined as early fibrosis, and stages 3-4 were defined as advanced fibrosis. Model 3 was established based on age and the four metabolite markers selected from Cohort 1 data using the RF model. It was then shown to successfully separate CLD patients with early fibrosis from those with advanced fibrosis in Cohort 1 with AUROC of 0.918 (0.889-0.946) and AUPR = 0.892 (0.854 - 0.925) (Figs. 4f, g). Model 3 results demonstrated better classification performances than those of APRI (AUROC=0.647, p<0.001), AST/ALT (AUROC=0.714, p<0.001) and FIB-4 (AUROC=0.802, p<0.001) in predicting liver fibrosis stages (Table 2). An example decision tree from Model 3 showed that the higher Tyr/Val ratio, Tyr, age and TCA indicated a higher risk of CLD with advanced firbosis (Fig. S5c).

A logit diagnostic RF-score for Model 3 differentiated CLD patients with early stage fibrosis from those with advanced fibrosis in Cohort 1 (Fig. 4j). The model yielded a sensitivity of 86.7% and specificity of 90.5% in Cohort 1 at a cutoff value of −0.115 (Table 2). After adjusting for HBV DNA levels, degree of necro-inflammation, HBeAb status, HBeAg status, liver function tests (i.e., PT, ALB, DBIL, IBIL), platelets, BMI, and medication (Entecavir) use, RF-scores remained statistically significant with a coefficient of 0.805 (p<0.001) (Table S3). The accuracy of our RF model was superior to those of AST/ALT ratio, APRI, and FIB-4 (Table 2 and Table S4).

### Validation of the predictive models in an independent HBV cohort (Cohort 2)

The metabolite markers identified and related models obtained in Cohort 1 were further validated for their liver fibrosis staging performance as well as for CLD diagnosis performance in Cohort 2, and the results were similar to those obtained from Cohort 1 (Table 2).

For the diagnosis of CLD patients, compared to APRI (AUROC = 0.879, AUPR = 0.958), AST/ALT (AUROC = 0.603, AUPR = 0.849), and FIB-4 (AUROC = 0.707, AUPR = 0.897), we again observed higher classification performances for Model 1 with AUROC of 0.977 (0.963-0.988) and AUPR of 0.993 (0.989-0.997) in the validation set (Figs. 3e, f). In addition, the Model 1 predicted RF-score in Cohort 2 differentiated CLD from NC with a sensitivity of 92.2% and specificity of 94.4% at the cutoff values determined for Cohort 1 (Fig. 3g, Table 2).

Applying Model 2 to Cohort 2 successfully discriminated cirrhotic patients from fibrotic patients with an AUROC of 0.844 (0.797-0.884) and AUPR of 0.827 (0.761-0.884) (Figs. 4c, d) and outperformed those of the APRI (AUROC=0.608, p<0.001), AST/ALT (AUROC=0.684, p<0.001) and FIB-4 (AUROC=0.758, p<0.001) indices. The Model 2 RF-score in Cohort 2 differentiated cirrhotic patients from fibrotic patients with a sensitivity of 71.8% and specificity of 81.6% at the same cutoff value used for the Cohort 1 data set (Fig. 4e, Table 2). Similarly, applying Model 3 to grade fibrosis stage in Cohort 2 resulted in greater performance with AUROC of 0.807 (0.756-0.852) and AUPR of 0.817 (0.764-0.866) (Figs. 4h, i) than those of APRI (AUROC=0.595, p<0.001), AST/ALT (AUROC=0.667, p<0.001) and FIB-4 (AUROC=0.739, p=0.01) indices. And the Model 3 RF-score also differentiated S0-2 fibrosis from S3-4 fibrosis with a sensitivity of 72.9% and specificity of 76.1% (Fig. 4j, Table 2).

We then introduced net reclassification improvement (NRI) and integral discriminant improvement (IDI) to quantify the improvement of our model to existing clinical indices. For different classification aims (Control vs. CLD, Fibrosis vs. Cirrhosis, S0-2 vs. S3-4) in an independent validation cohort (Cohort 2), the categorical and the continuous NRI and IDI of the RF models all achieved positive values when compared to FIB-4, APRI and AST/ALT, suggesting an augmentation of classification performances for our biomarker panel and RF models (Table S5).

### Classification of S0-2 vs. S3 vs. S4

In addition to the binary classifications that we have performed, we further determined whether our biomarker panel could classify multiple groups among CLD patients. We trained a new RF model with the marker panel and applied multinomial regression to APRI, AST/ALT and FIB-4 respectively for the discrimination of S0-2 vs. S3 vs. S4 using Cohort 1. We compared their performances on Cohort 1 (OOB predictions of RF model) and Cohort 2 using micro-average AUROC and AUPR and we found that our marker panel-based multi-group classifier outperformed other methods. In the Cohort 1 data, our classifier showed higher AUROC of 0.944 (0.928-0.963) and AUPR of 0.908 (0.883-0.938) compared to APRI (AUROC=0.79, AUPR=0.658), AST/ALT (AUROC=0.817, AUPR=0.688), and FIB-4 (AUROC=0.858, AUPR=0.774) (Fig. S6a, b). In the Cohort 2 validation data, our marker panel classifier consistently displayed higher AUROC of 0.841 (0.799-0.885) and AUPR of 0.748 (0.674-0.81) compared to APRI (AUROC=0.790, AUPR=0.608), AST/ALT (AUROC=0.772, AUPR=0.597), and FIB-4 (AUROC=0.816, AUPR=0.699) (Figs. S6c, d).

## DISCUSSION

As the prevalence of CLD rises worldwide, accurate and reliable assessments for the severity of this disease are increasingly important for treatment selection and longitudinal monitoring [13]. Attempts to develop noninvasive tools for staging CLD have yielded multiple scores, indices, and imaging modalities [4-7, 10] that might be used *in lieu* of liver biopsy, with the AST/ALT ratio, APRI, and FIB-4 as examples [5-7]. Current noninvasive assessments have the advantage of allowing repeated applications and are well-received by the patients. In this study, we identified a panel of metabolite markers that consisted of C18:2 n6t, TCA, Tyr, and a Tyr/Val ratio, that was highly correlated with discrete stages of CLD progression in patients with HBV infection.

Histologic staging of CLD by liver biopsy provided a reference standard for our study. In the Scheuer system, one of the most clinically validated systems for staging liver fibrosis, S0 is defined as no fibrosis, S1 as portal fibrosis, S2 as periportal fibrosis, S3 as septal fibrosis, and S4 as cirrhosis [38]. The clinically overt stage of cirrhosis includes compensated cirrhosis with/without portal hypertension and decompensated cirrhosis [39]. In this study, we first identified candidate markers that significantly differed between NC and patients with CLD that correlated well with fibrotic stage and necro-inflammation based on univariate, LASSO and RF analyses. We then constructed diagnostic models to discriminate CLD patients from NC, and to discriminate CLD patients at different fibrosis stages, i.e., early vs. advanced fibrosis (S0-2 *vs*. S3-4) and fibrosis *vs*. cirrhosis (S0-3 *vs*. S4). This resulted in three optimized marker panel-based RF predictive models for staging liver fibrosis that, upon validation, showed acceptable performance across independent cohort. The AUROC and AUPR of our biomarker panel were significantly greater than those of the AST/ALT ratio, APRI and FIB-4, suggesting superior predictive value for this metabolite marker panel.

Altered BA profile and BA synthesis are associated with various hepatic diseases, such as chronic hepatitis B, primary biliary cirrhosis, chronic hepatitis C, and NAFLD. Circulating BAs are commonly used in clinical practice to assist evaluation of the severity of CLD [40, 41]. Several studies, including our previous work, on cirrhosis and HCC have shown dramatically increased levels of GCA, GCDCA, TCA, and TCDCA in the circulation of patients with NAFLD [42], NASH [42], HBV [43], cirrhosis [44], and HCC [44]. The liver also plays a major role in lipid metabolism by taking up FFAs, and manufacturing, storing, and transporting lipid metabolites [45, 46]. A characteristic pattern of plasma amino acids has been described in cirrhotic subjects [47-49] and in samples collected in England and the USA, metabolic and biochemical differences have been shown between stable and unstable cirrhotics [47, 48]. Advanced liver fibrosis, especially cirrhosis, was also associated with altered plasma AA patterns, including decreased levels of branched chain amino acids (leucine, isoleucine, valine) and increased concentrations of the aromatic amino acids phenylalanine and tyrosine [21]. An index based on AA concentration has already been proposed for diagnosing liver fibrosis [22]. In patients admitted to either the Veterans Administration Hospital or the Yale-New Haven Medical Center between 1 January 1965 and 1 May 1966, fasting tyrosine levels tended to be slightly increased in patients with hepatitis and markedly increased in patients with cirrhosis [50]. The present study showed that a combined panel of FFA, BA, and AA was a strong predictor for CLD progress.

Linoelaidic acid is an isomer of linoleic acid. It has been reported that linoelaidic acid may inhibit the development of tumors through its antioxidant effects, has a role in the prevention of atherosclerosis and modulates certain aspects of immune system [51]. The significantly decreased levels of linoelaidic acid may thus be an indication of a disease state. Further research on these findings and human epidemiological data is warranted to confirm this.

The major strengths of our study were the use of large sample sizes to construct and verify all models, and the quantification of the metabolite markers (BA, FFA and AA) using standardized protocols. Furthermore, participants in the validation set (Cohort 2) were recruited independently from those in Cohort 1, and this new set of patients confirmed the robustness of our marker panel and predictive models.

The limitations of our study included: (1) Use of medications was a confounding factor for our model but key findings were not altered after correcting for medication use. Larger studies are needed to further evaluate the effect of these medications; (2) HBV infection was the only or major cause of CLD in this study, and the participants were all Chinese. Therefore, the results may not be extrapolated to CLD with other etiologies outside these diseases, or to other racial/ethnic groups. Future large-scale validation studies should include CLD with other etiologies, and participants of other race/ethnicity, before implementing this 4-marker panel in clinical practice; (3) In addition to cross-sectional studies, longitudinal studies are needed to further validate the reproducibility of the current findings and the predictive values of the models, especially those used to differentiate early from advanced liver fibrosis; and (4) The cost of full spectrum metabolomic analysis is high. However, if the robustness of this 4-marker panel is proven in future validation studies, specific tests may be developed for only C18:2 n6t, TCA, Tyr, and Val to decrease the cost, and to translate this marker panel to clinical practice.

## Conclusions

In summary, using targeted metabolomics analyses, we identified four metabolite markers from serum that accurately differentiated CLD patients from NC, and differentiated varied stages of liver fibrosis, including S0-2 vs. S3-4, and S0-3 vs. S4. The diagnostic performance of this novel, noninvasive 4-marker panel was superior to FIB-4, AST/ALT ratio, and APRI. If validated in future studies, this 4-marker panel will be useful in reducing the need for liver biopsies in identifying patients with non-significant fibrosis, as well as aiding in the continued assessment of CLD in patients previously diagnosed with CLD.

## ACKNOWLEDGMENTS

We wish to thank the research coordinators of the participating hospitals for their assistance in collecting clinical data and samples.

## Abbreviations

CLD: chronic liver disease
ROC: receiver operating characteristic
PR: precision-recall
AUROC: area under the ROC curve
BAs: bile acids
FFAs: free fatty acids
AAs: amino acids
HBV: hepatitis B virus
Cis: confidence intervals
C18:2 n6t: linolelaidic acid
TCA: taurocholate
tyr: tyrosine
Val: valine
ALT: alanine transaminase
AST: aspartate transaminase
TBIL: total bilirubin
ALP: alkaline phosphatase
GGT: gamma-glutamyl transferase
ALB: Albumin
PALB: prealbumin
TBA: Total bile acid
CHE: cholinesterase
CREA: creatinine
BUN: blood urea nitrogen
CHOL: cholesterol
TG: Triglyceride
HDLC: High-Density Lipoprotein Cholesterol
LDLC: low-density lipoprotein cholesterol
ApoAI: Apolipoprotein A1
ApoB: Apolipoprotein B
PT: Prothrombin Time
Fib: Fibrinogen
GLU: Glucose
RBC: red blood cell count
WBC: white blood cell
HCT: hematocrit
HGB: hemoglobin
MCH: mean corpuscular hemoglobin
MCHC: Mean corpuscular hemoglobin concentration
MPV: Mean platelet volume
PLT: platelet
GLB: globin

## Notes

**Financial support:** This study was financially supported by the National Key R&D Program of China (2017YFC0906800), the National Institutes of Health/National Cancer Institute Grant 1U01CA188387-01A1, the National Natural Science Foundation of China (81160439, 81573873), the National Key R&D Program of China (2017YFC0906800), the Natural Science Foundation of Shanghai, China (14ZR1441400, 12ZR1432300), China Postdoctoral Science Foundation funded project, China (2016T90381, 2015M581652), and E-institutes of Shanghai Municipal Education Commission, China (E03008, 085ZY1205). The funding sources did not have any role in the design and conduct of the study; collection, management, analysis, and interpretation of the data; preparation, review, or approval of the manuscript; and decision to submit the manuscript for publication.

**DISCLOSURES:** All authors read and approved the final manuscript and declared no conflicts of interest.

### Competing Interest Statement

The authors have declared no competing interest.

### Funding Statement

This study was financially supported by the National Key R&D Program of China (2017YFC0906800), the National Institutes of Health/National Cancer Institute Grant 1U01CA188387-01A1, the National Natural Science Foundation of China (81160439, 81573873), the National Key R&D Program of China (2017YFC0906800), the Natural Science Foundation of Shanghai, China (14ZR1441400, 12ZR1432300), China Postdoctoral Science Foundation funded project, China (2016T90381, 2015M581652), and E-institutes of Shanghai Municipal Education Commission, China (E03008, 085ZY1205). The funding sources did not have any role in the design and conduct of the study; collection, management, analysis, and interpretation of the data; preparation, review, or approval of the manuscript; and decision to submit the manuscript for publication.

